# Radiographic classification of mandibular osteoradionecrosis: A blinded prospective multi-disciplinary interobserver diagnostic performance study

**DOI:** 10.1101/2025.02.11.25322082

## Abstract

**Background:** Osteoradionecrosis of the jaw (ORNJ) is a debilitating complication that affects up to 15% of head and neck cancer patients who undergo radiotherapy. The ASCO/ISOO/MASCC-endorsed ClinRad severity classification system was recently proposed (and recommended in the latest ASCO guidelines) to incorporate radiographic findings for determining ORNJ severity based on the vertical extent of bone necrosis. However, variability in imaging modalities and specialty-specific knowledge may contribute to disparities in diagnosing and classifying ORNJ. This study aims to evaluate and benchmark multi-specialty physician performance in diagnosing and severity classification of ORNJ using different radiographic imaging.

**Methods:** A single institution retrospective diagnostic validation study was conducted at The University of Texas MD Anderson Cancer Center involving 20 healthcare providers across varying specialties including oral oncology, radiation oncology, surgery, and neuroradiology. Participants reviewed 85 de-identified imaging sets including computed tomography (CT) and orthopantomogram (OPG) images from 30 patients with confirmed ORN, with blinded replicates (n=10) for assessment of intra-observer variability and asked to diagnose and stage ORNJ using the ClinRad system. Diagnostic performance was assessed using ROC curves; intra- and inter-observer agreement were measured with Cohen’s and Fleiss kappa, respectively. Sub-analyses considered physician specialty, years of clinical experience and level of confidence.

**Results:** Paired CT-OPG imaging improved ORNJ diagnostic performance across all specialties, with AUC values ranging from 0.79 (residents) to 0.98 (surgeons). Inter- and intra-rater agreements for ORNJ detection were limited, with median (IQR) Fleiss and Cohen’s kappa values of 0.38 (0.22) and 0.08 (0.17), respectively. Slight to fair inter-rater agreement in severity classification ORNJ was observed with median (IQR) Fleiss kappa values of 0.22, 0.13, and 0.05 for stages 0/1, 2, and 3, respectively. The most commonly reported radiographic features for confirmed ORNJ cases staged as ClinRad grade 1 or 2 were “bone necrosis confined to alveolar bone” (22.7%), “bone necrosis involving the basilar bone or maxillary sinus” (14.8%), and “bone lysis/sclerosis” (20.0%).

**Conclusion:** This study establishes an essential benchmark for physician detection of radiographic ORNJ. The significant variability in diagnostic and severity classification observed across specialties emphasizes the need for standardized imaging protocols and specialist training as well as highlights the value of multimodality imaging.

## 1. Introduction

Globally, head and neck cancers (HNC) rank as the seventh most prevalent cancer, affecting upwards of 890,000 individuals annually worldwide and rising^1^. This rise is especially notable in oropharyngeal cancers (OPC) associated with the human papillomavirus (HPV)^2,3^, generally affecting a younger population who are healthier and typically present with better dentition compared to older HNC populations due to factors such as poor dental health and smoking. In the United States alone, radiation therapy (RT) serves as a critical treatment for over 63,000 individuals annually, especially for HPV-associated (HPV+) cancers^4^. With a longer life expectancy, oropharyngeal cancer (OPC) survivors are at a higher risk of developing long-term RT-induced side effects such as osteoradionecrosis of the jaw (ORNJ), a highly debilitating sequalae developed in 4-15%^4^ of patients with HNC. This morbid complication not only greatly affects patients’ quality of life (QoL) but also often results in high medical costs^5^.

ORNJ manifests in the mandible as a state of devitalized bone with inadequate healing over time due to direct cellular damage and vascular injury from radiation, resulting in reduced blood supply and subsequent bone death (necrosis)^6^. The composition and density of the mandible bone varies, with two defined regions: the more porous alveolar region surrounding the teeth and the denser basal region extending beyond the alveolar canal. The extent of necrotic bone can range from the surface of the alveolar region to beyond the alveolar bone and into the basal bone region. Bone death can be identified clinically (i.e., exposed bone) and/or radiographically (i.e., sclerosis or pathologic fracture), and diagnostic imaging is key in spatially identifying the extent of these anatomical changes. Existing physician-utilized grading systems for severity classification of ORN has predominantly relied on patient symptoms and oral clinical assessments. The ClinRad severity classification system was recently proposed^7^ (and recommended in the latest ASCO guidelines^8^) to incorporate radiographic findings for determining ORNJ severity based on the vertical extent of bone necrosis (as well as presence/absence of exposed bone or fistula), which is particularly relevant in diagnosing early ORNJ cases with intact mucosa. Moreover, a recent Delphi study^9^ provided strong consensus on the importance of including radiographic findings in a formal definition of ORN. However, there is still an unmet need for standardization of these image-based features.

Diagnostic imaging methods typically vary across the different specialties involved in the management of ORN. Dental specialists routinely rely on panoramic imaging such as orthopantomogram (OPG), due to its cost-effectiveness, accessibility, and ability to provide a broad overview of the dental structures. Conversely, radiologists and oncologists tend to use computed tomography (CT) images, which offer superior spatial resolution and detailed visualization of bony structures, making it particularly valuable for detecting subtle changes in bone density and identifying more advanced ORNJ stages. The variability in imaging modalities combined with the subjectivity of clinician-based diagnosis and the use of multiple severity classification systems, contributes to disparities in detection and classification across institutions and specialists. Additionally, emerging imaging techniques, including MRI sequences like dynamic contrast-enhanced^10^ and/or black bone imaging^11^, are increasingly being studied in research contexts and may hold promise for further advancing diagnostic precision in the future. However, at present, volumetric CT and OPG remain the primary imaging modalities in clinical practice, aligning with the focus of this study.

Previous works^10,12,13^ have shown that computer-based approaches may enable earlier detection of quantitative density changes subsequently being more objective than clinician-based diagnosis and staging. However, to develop a benchmark for computer-based approaches, we sought to establish the limitations and baseline capabilities of clinical radiographic review, in order to establish “ground truth” diagnostic accuracy for ORNJ.

In this study, we aimed to i) benchmark clinician performance in the radiographic detection and severity classification of ORNJ across various specialties, providing an essential reference for diagnostic accuracy using the ASCO/ISOO/MASCC-endorsed ClinRad^7^ system, ii) compare the effectiveness of CT and OPG in ORNJ detection, and iii) generate testable hypotheses for future research, laying the groundwork for studies aimed at refining diagnostic criteria and exploring advanced computational tools to enhance consistency and accuracy in ORNJ identification.

## 2. Materials and Methods

### 2.1 Study design

This study was designed as a single institution retrospective diagnostic validation study at The University of Texas MD Anderson Cancer Center (MD Anderson) under the ethical approval of the RCR-03-0800 protocol, which includes a waiver of informed consent for image analysis and segmentation tasks. As such, individual informed consent was not required, as the study involved retrospective diagnostic validation of anonymized images without patient interaction. The research adhered to institutional guidelines, the principles outlined in the Declaration of Helsinki, and good clinical practice (GCP) standards. Analysis was performed as per CROSS (Checklist for Reporting of Survey Studies)^14^ and the STARD for Abstracts (Checklist for reporting diagnostic accuracy)^15^ guidelines (see Supplement A). The target population for this questionnaire included physicians from varying levels of clinical experience and specialties involved in the diagnosis and management of ORN, such as dentists, radiation oncologists, surgeons, neuroradiologists, and residents.

### 2.2 Patient selection

Patients were retrospectively selected from the clinical database maintained at MD Anderson based on inclusion criteria of diagnosed ORNJ post RT confirmed through clinical charts. All patients included in this study had been clinically diagnosed with mandibular ORN; patients without definitive ORNJ diagnosis (n=30), with ORNJ in locations other than the mandible (e.g., maxilla) (n=2) or with incomplete imaging data were excluded. A flow diagram of inclusion and exclusion diagram can be found in Supplement C. A summary of the demographic and clinical characteristics of the included patient cohort is presented in Supplement D.

### 2.3 Sample Size

Following the guidelines provided by Gwet et al.,^16,17^ pre-testing minimum required sample size for required participants/cases was calculated as per Equation 1, assuming a 30% relative inter-observer error, an anticipated overall inter-observer agreement probability of 0.61 (modest agreement threshold)^18^, our calculations indicated that a minimum of 55 cases/image sets would be necessary to ensure the reliability of our kappa estimates at a significance level of α=0.05 with 20 physician raters.

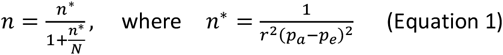

### 2.3 ORNJ stage definition

The ClinRad severity classification system^7^ defines four stages of ORNJ (see supplementary Table E1): The pre-ORNJ stage, referred to as minor bone spicules, involves superficial mobile spicules or sequestra within the mucosa without any radiographic evidence of bone necrosis; Stage 0 includes radiographic evidence of bone necrosis confined to the alveolar bone, with features such as bone lysis, sclerosis, widening of the periodontal ligament space, or absence of osseous filling of extraction sockets, but with intact mucosa and no clinical signs of ORN; Stage 1 involves clinical signs of ORN, such as exposed bone or intact mucosa if confined to the alveolar bone, with or without radiographic evidence; Stage 2 is characterized by radiographic evidence involving the basilar bone or maxillary sinus, which may present with exposed bone or intact mucosa; finally, Stage 3 represents advanced ORN, presenting with severe complications, such as pathologic fractures, orocutaneous fistulas, or oral antral/oral nasal communication. For this study we only considered stages that could be radiographically assessed i.e. stages 0/1 (as one stage), 2, and 3.

### 2.4 Image data

Physicians examined 85 imaging studies, including head and neck CT and OPG images obtained at pre-RT and post-ORNJ time points for a total of 30 patients. All patients included in this study eventually developed ORNJ; however, their pre-RT and pre-ORNJ scans were used as control cases for comparison. Not all patients had OPG images obtained before or after ORNJ diagnosis; only 9 subjects out of the 30 subjects had usable OPGs. Thus, up to four imaging studies were selected for each patient (Figure 1), including two CT and two OPG images taken both pre-RT and post-ORNJ diagnosis. The two image modalities, CT and OPG, were first presented independently for all patients as individual cases; then, for a subset of patients, paired CT and OPG image sets were presented again, and reviewers were asked to repeat the diagnosis and severity classification exercises. Ten patients were then used as duplicates and their images were presented to the reviewers as blinded repeats to assess intra-observer variability.

**Figure 1.**
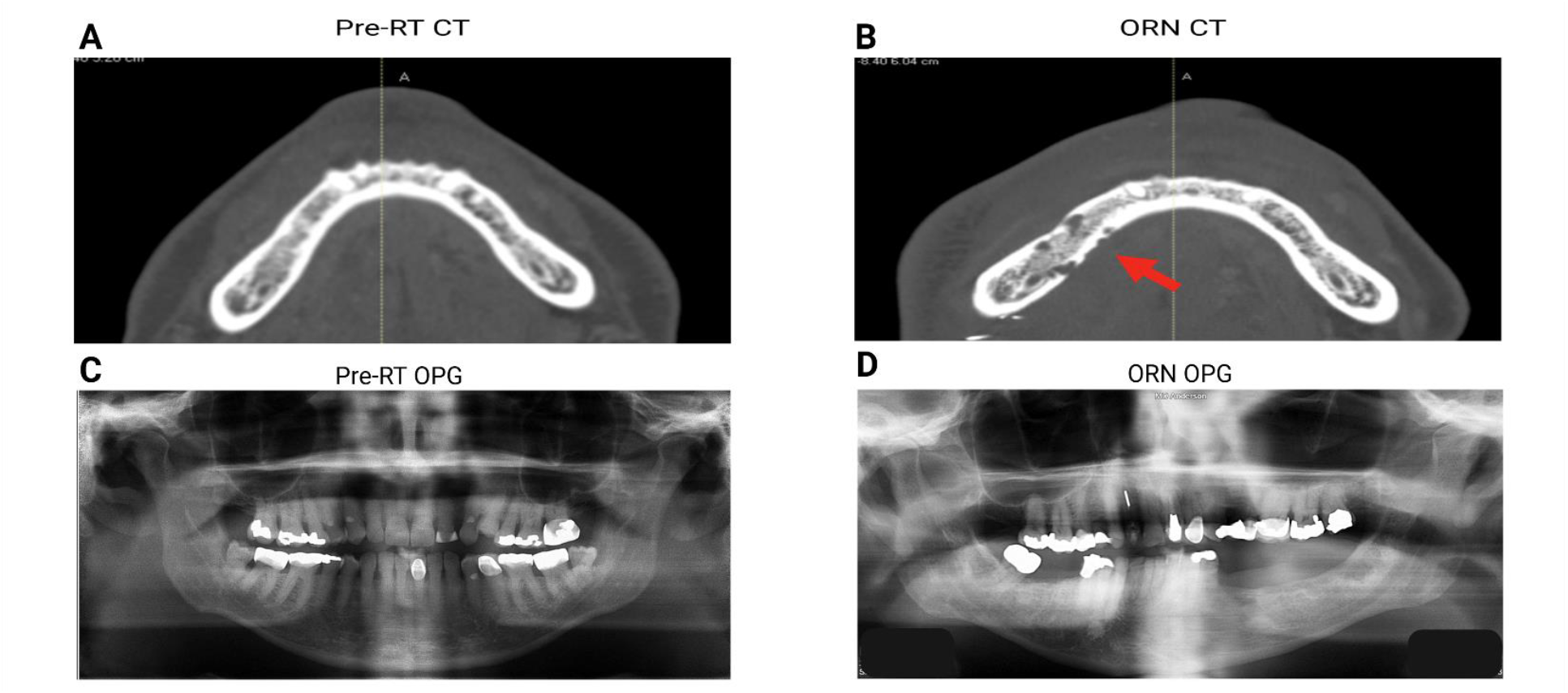
Examples of pre- and post-RT imaging modalities. Panels A and C show axial computerized tomography (CT) and orthopantomograms (OPG) scans of the mandible pre-RT, respectively. Panels B and D display corresponding post-RT images, in this case for a patient who developed ORN. These images represent the dataset provided to survey participants for the ORNJ diagnosis and severity classification assessments.

The “ground truth” for ORNJ status was derived from clinical charts, including the date of ORNJ diagnosis and the start date of RT. Radiographic imaging sets were retrieved from Epic (Verona, Wisconsin), a widely used electronic health record system that integrates patient information, medical imaging, and clinical workflows, via an integrated picture and archiving computer systems (PACS). CT images were uploaded into a commercial research treatment planning system (TPS; RayStation v12 A, RaySearch Laboratories, Stockholm, SE) whereas OPG (2D) images were viewed independently as 2D-minensional image captures (Joint Photographic Experts Group, JPEG). To further standardize imaging parameters, acquisition metadata were extracted from the DICOM headers, including tube voltage (kVp), slice thickness, exposure time, x-ray tube current, and spatial resolution details. Complete CT acquisition parameter details are provided in Supplement F.

To maintain blinding, all subjects were anonymized, removing name, sex, date of birth, and given a non-sequential subject ID. They were then shuffled and randomly assigned a blinded study ID to minimize bias. Window-level default settings were standardized across all images (“Bone Window”, W1500-L300) to ensure consistent display and contrast of relevant anatomical structures across all images at initial display (although readers could re-window/re-level at their discretion). In-person review was performed under uniform lighting conditions in natural light conditions (n=18); however, owing to mobility/availability, 2 readers undertook remote review.

### 2.5 Survey and data collection

Physicians who treat or diagnose ORNJ at our facility were recruited via email with an invitation to join the study and notified that two in-person sessions over three hours each were required. A 20-minute onboarding was conducted to show participants the ClinRad system in the context of this study (e.g., supplementary Figure E1), the RayStation TPS, and a facilitated discussion on radiographic ORNJ presentation in CT. Then, physicians completed a Qualtrics survey consisting of three sections: a pre-survey, a main survey, and a paired survey (see Supplement G). The pre-survey collected participants’ background data, including years of experience, familiarity with the index commercial TPS interface, and the approximate number of ORNJ cases observed annually. The main survey was designed to capture their diagnostic and severity classification decisions, the decision-making process (including identification of observed anatomical abnormalities), and confidence levels in each assessment using a Likert scale. The paired survey assessed diagnostic consistency using combined 2D/3D imaging sets (i.e. OPG and CT). All participants were asked to complete the main survey for all cases, and these cases were provided in a randomized order. To identify each participant, they were given a unique *reader ID*; similarly, each image/case/subject had a unique *subject ID*.

### 2.6 Statistical analyses

The primary outputs of this study included ORNJ diagnosis (ORNJ vs. non-ORN), ORNJ severity classification using the ClinRad system with associated radiographic findings, and confidence levels for diagnostic and severity classification decisions. Sensitivity and accuracy were analyzed using ROC curves across imaging modalities (CT, OPG, and paired CT-OPG), with sensitivity, specificity, and AUC values compared by specialty, years of experience, and annual ORNJ cases managed using the DeLong’s statistical test; additionally, results were compared to chance. AUC was computed using a binary classification approach, where predictions were assessed based on whether physicians correctly identified clinically diagnosed ORNJ cases. Confidence scores were not incorporated, as the analysis relied solely on binary correct/incorrect classification outcomes rather than probability estimates per prediction. The Fleiss Kappa statistic, specifically designed for assessing agreement between multiple raters when the raters are assigning categorical ratings to a number of items^17,19^, was applied to quantify the degree of consistency and agreement among physicians in identifying and classifying severity of ORNJ based on radiographic information only and to highlight potential systematic differences between specialties. Fleiss Kappa threshold values were used to indicate slight (0.0-0.20), fair (0.21-0.40), moderate (0.41-0.60), substantial (0.61-0.80) or almost perfect (0.81-1.00) agreement^18^. Single-modality and paired-modality results were compared to identify systematic differences across specialties as well as to assess the potential benefits of combining image modalities. For intra-rater variability, Cohen’s Kappa statistic was applied to assess the consistency of individual physicians in detecting and reporting severity classification of ORNJ across repeated evaluations of the same cases. Threshold values similar to those used for Fleiss Kappa were applied to interpret the agreement levels. To account for missing data, case-specific calculations were performed for ORNJ detection, staging, radiographic findings, and Fleiss Kappa analyses, ensuring a robust evaluation of the reliability of radiographic ORNJ detection and severity classification using the ClinRad system.

## 3. Results

### 3.1 Participants

Responses were collected from 20 observers, including 5 radiation oncology residents and one general practice resident (30%)—4 surgeons (20%), 4 radiation oncologists (20%), 3 dentists (15%), and 3 neuroradiologists (15%), with the highest response rate (23%) obtained from surgeons, oncologists, and residents (see Table 1), where a fully completed survey consisted of 105 responses per participant. Out of the total 85 images, 45 were taken prior to the ORNJ diagnosis, while 40 were with active or pot-treatment ORNJ.

**Table 1.**
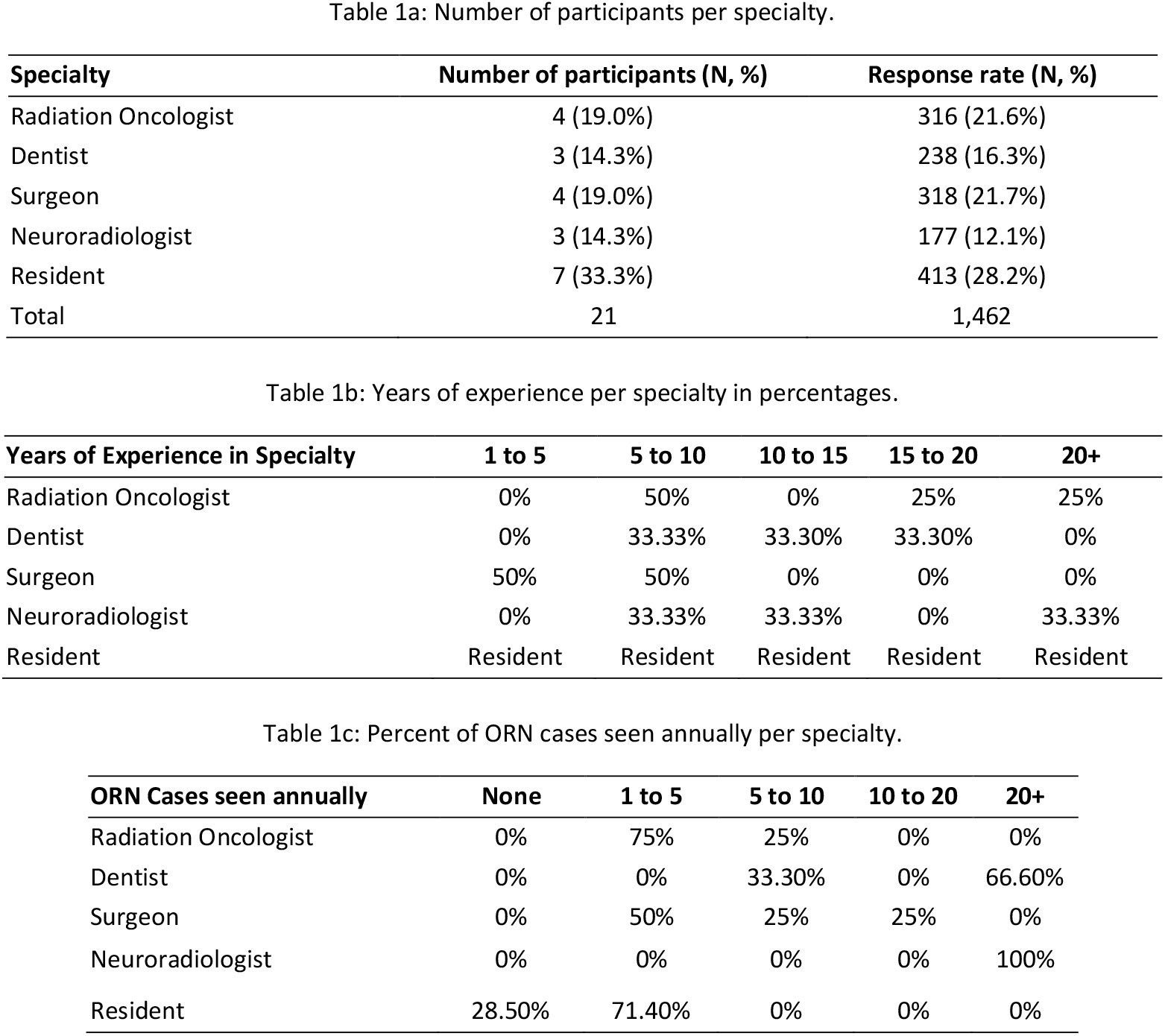
Descriptive characteristics of the survey participants cohort (additional related tables are provided in Supplement B)

### 3.2 ORNJ Detection

ROC analysis revealed variability in ORNJ diagnostic accuracy by specialty and years of experience (see Supplement H). Non-paired assessments using CT alone showed varying accuracy across specialties and experience levels; dentists with 10–15 years of experience achieved 61.3% ± 7.2% accuracy, while radiation oncologists with 15–20 years reached 55.2% ± 2.1%. Surgeons with 5–10 years achieved 54.7% ± 8.9% accuracy, but their sensitivity and specificity varied widely, ranging from 59.4% ± 4.5% to 48.5% ± 4.5% respectively. Neuroradiologists with 20+ years showed high sensitivity on OPG imaging (100% ± 30.7%) but low specificity (30% ± 41.7%).

The general practitioner resident demonstrated high sensitivity on CT (87.5% ± 29.7%) but low specificity (14.7% ± 29.7%), indicating a tendency for over-diagnosis. For OPG-only assessments, accuracy across specialties declined, with radiation oncology residents achieving 28.6% ± 10.1% and neuroradiologists with 10–15 years achieving 37.5% ± 15.4%. Paired CT-OPG imaging (Figure 2) improved accuracy across all specialties, with AUC values ranging from 79% [CI: 0.68, 0.82] (residents) to 98% [CI: 0.92, 0.99] (surgeons), emphasizing the benefit of multimodal imaging for ORNJ detection.

**Figure 2.**
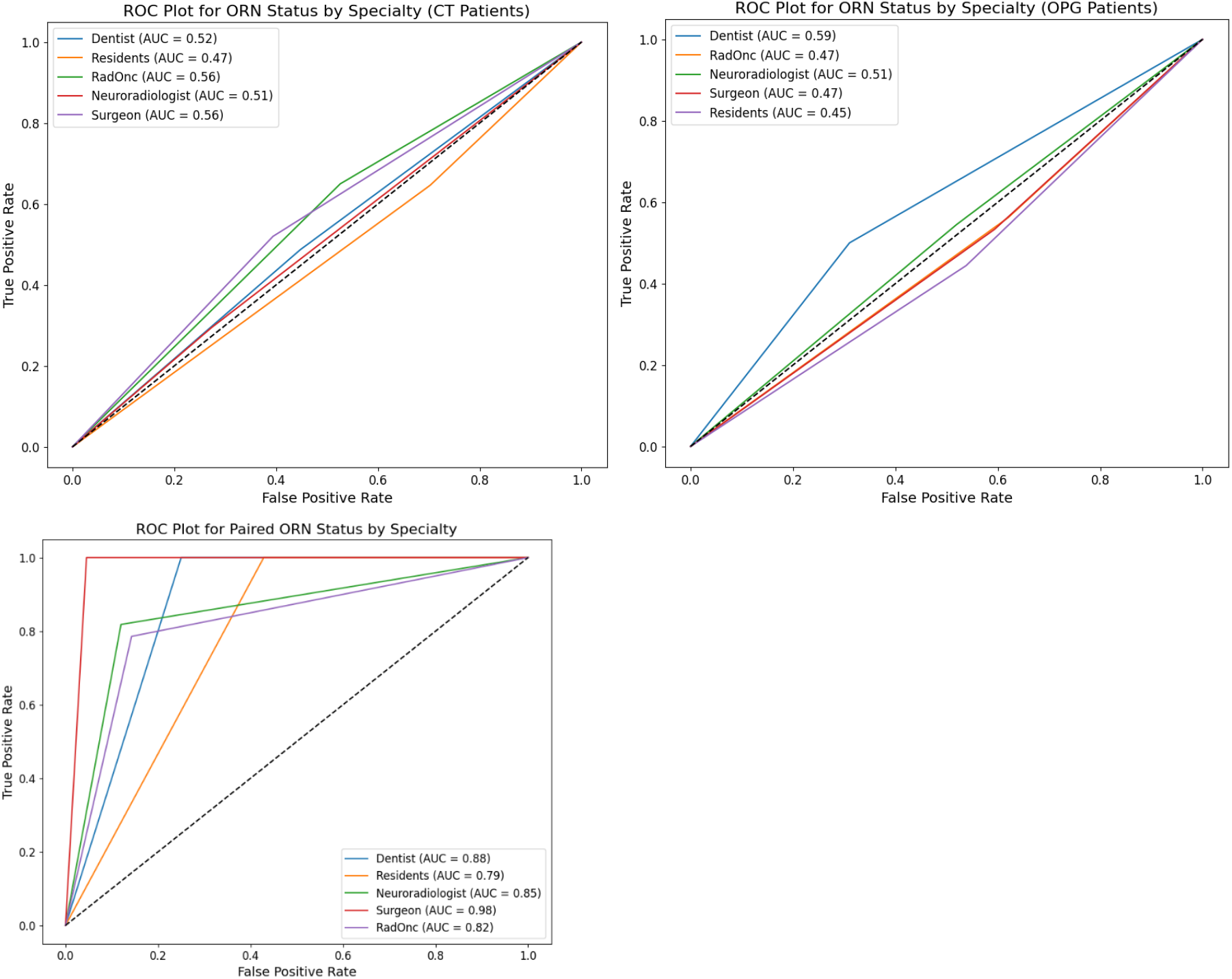
ROC curves illustrate the diagnostic performance of different clinical specialties in detecting ORNJ status on CT (top left), OPG (top right) and paired (bottom) images.

Statistically significant of differences in AUC values across specialties was assessed using the DeLong test (Supplement I). In particular, the general practitioner resident demonstrated statistically significantly lower AUC values compared to neuroradiologists and radiation oncologists (DeLong’s test p < 0.0001), suggesting reduced ability to detect ORN on CT. Neuroradiologists exhibited higher AUCs than dentists and radiation oncology residents (p < 0.0001). In contrast, surgeons and radiation oncologists showed no significant differences (p > 0.05), indicating comparable diagnostic performance. Despite these differences, AUC values across all specialties remained relatively low, highlighting the overall difficulty in radiographically detecting ORN.

Additional DeLong analyses comparing specialty AUCs against chance (AUC = 0.5) revealed that none of the specialties demonstrated statistically significant deviation from random classification after Bonferroni correction (p > 0.05 for all comparisons, supplementary Table I2). Although radiation oncologists had the highest uncorrected AUC (0.552, p = 0.062), this result was not statistically significant after multiple testing correction, indicating that their performance did not significantly exceed chance levels. Similarly, dentists (AUC = 0.549, p = 0.133) and surgeons (AUC = 0.519, p = 0.501) showed slightly above-chance performance, but these differences also failed to reach statistical significance. The general practitioner resident and neuroradiologists both had AUC values of 0.500, demonstrating no apparent ability to differentiate between ORN and non-ORN cases beyond chance. Radiation oncology residents exhibited the lowest AUC (0.456, p = 0.184), suggesting a potential tendency toward misclassification. Collectively, these findings indicate that, despite observed differences between specialties, no group demonstrated a statistically significant ability to detect ORN beyond chance, reinforcing the inherent difficulty of ORN diagnosis based on radiographic imaging alone. These results highlight the need for improved imaging-based diagnostic strategies, including advanced imaging modalities or decision-support tools, to enhance ORN detection across specialties. When using paired 2D/3D imaging, all specialties demonstrated significantly improved AUC values (p < 0.001 for all, supplementary Table H3), with surgeons achieving the highest AUC (0.977) and residents the lowest (0.786). Unlike the single-modality results (supplementary tables H1 and H2), all specialties showed performance significantly exceeding chance (p < 0.001). However, DeLong’s test revealed no significant differences between specialties (p > 0.05 for all comparisons), suggesting that paired imaging benefits all groups equally.

### 3.3 ORNJ Staging

A number of non-ORNJ cases were classified as Stage I across specialties, with the general practitioner resident showing the highest misclassification rate (72.1%), followed by neuroradiologists with 20+ years of experience (60%) and surgeons with 5-10 years of experience (44.2%), Similarly, radiation oncology residents misclassified 35% of non-ORNJ cases as Stage I, indicating a need for improved diagnostic precision across experience levels.

Of the confirmed ORNJ cases, most were staged as Stage I across all modalities with the general practitioner resident (67.6%) and radiation oncologists with 15-20 years of experience (52.5%) showing the highest rates of classifying ORNJ cases as Stage I. Notably, neuroradiologists with 20+ years of experience classified 80% of ORNJ cases as Stage I on OPG imaging, indicating potential overconfidence or misidentification in advanced stages (Figure 3). Confidence levels for severity classification (supplementary Figure J1) varied significantly across specialties and modalities, with lower confidence (levels 2 and 3) predominantly associated with non-ORNJ and Stage I cases, while higher confidence (levels 4 and 5) was more frequently observed in correctly staged advanced ORNJ cases. This trend highlights variability in diagnostic confidence, particularly among neuroradiologists, across all imaging modalities.

**Figure 3.**
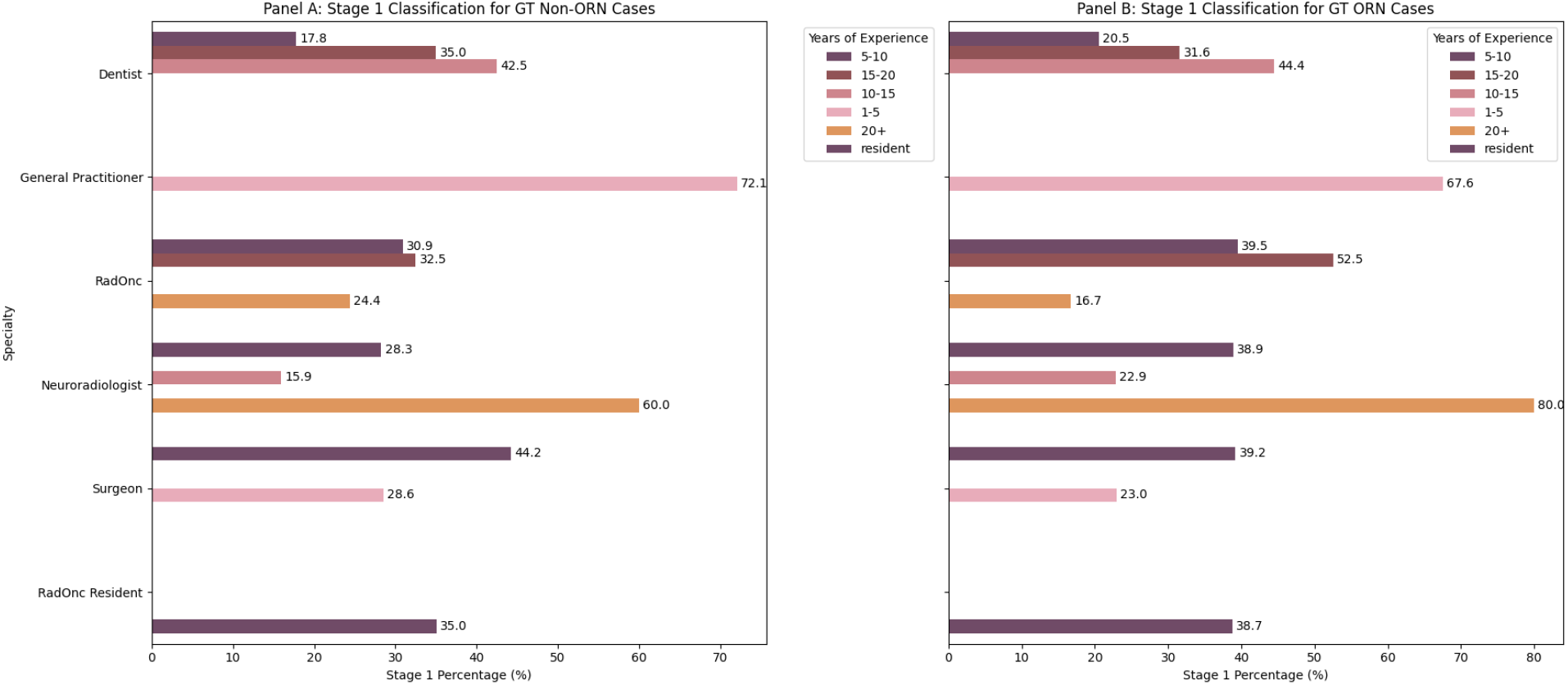
Panel A (left) displays the percentage of ground truth (GT) non-ORNJ cases that were misclassified as Stage I by specialty and years of experience. Panel B (right) shows the percentage of GT ORNJ cases that were classified as Stage I across the same categories. The bars represent the distribution of Stage I classifications, segmented by the different specialties and levels of experience.

### 3.4 Radiographic findings

Figure 4 summarizes the radiographic features by ORNJ stage as determined by survey participants. Among ORNJ cases (i.e., ORNJ positive ground truth determined from clinical reports), the most frequently selected features were “Bone necrosis confined to alveolar bone” (22.7%), “Bone lysis/sclerosis” (20.0%), and “Basilar bone involvement” (14.8%), while 20.7% showed no radiographic findings. For ground truth non-ORNJ cases, the most common features were no radiographic findings (23.7%), followed by “Alveolar bone necrosis” (19.9%) and “Bone lysis/sclerosis” (18.3%). Stage-specific trends showed that Stage 1 was primarily associated with “Alveolar bone necrosis” (26.9%) and “Bone lysis/sclerosis” (21.0%). Stage 2 cases had increased selection of “Basilar bone involvement” (14.8%) and “Bone lysis/sclerosis” (20.0%). In Stage 3, the most common findings were “Pathological fracture” (26.6%) and “Basilar bone involvement” (20.9%). Misidentified ORNJ cases showed a high occurrence of “No radiographic findings” (23.7%), “Alveolar bone necrosis” (19.9%), and “Bone lysis/sclerosis” (18.3%).

**Figure 4.**
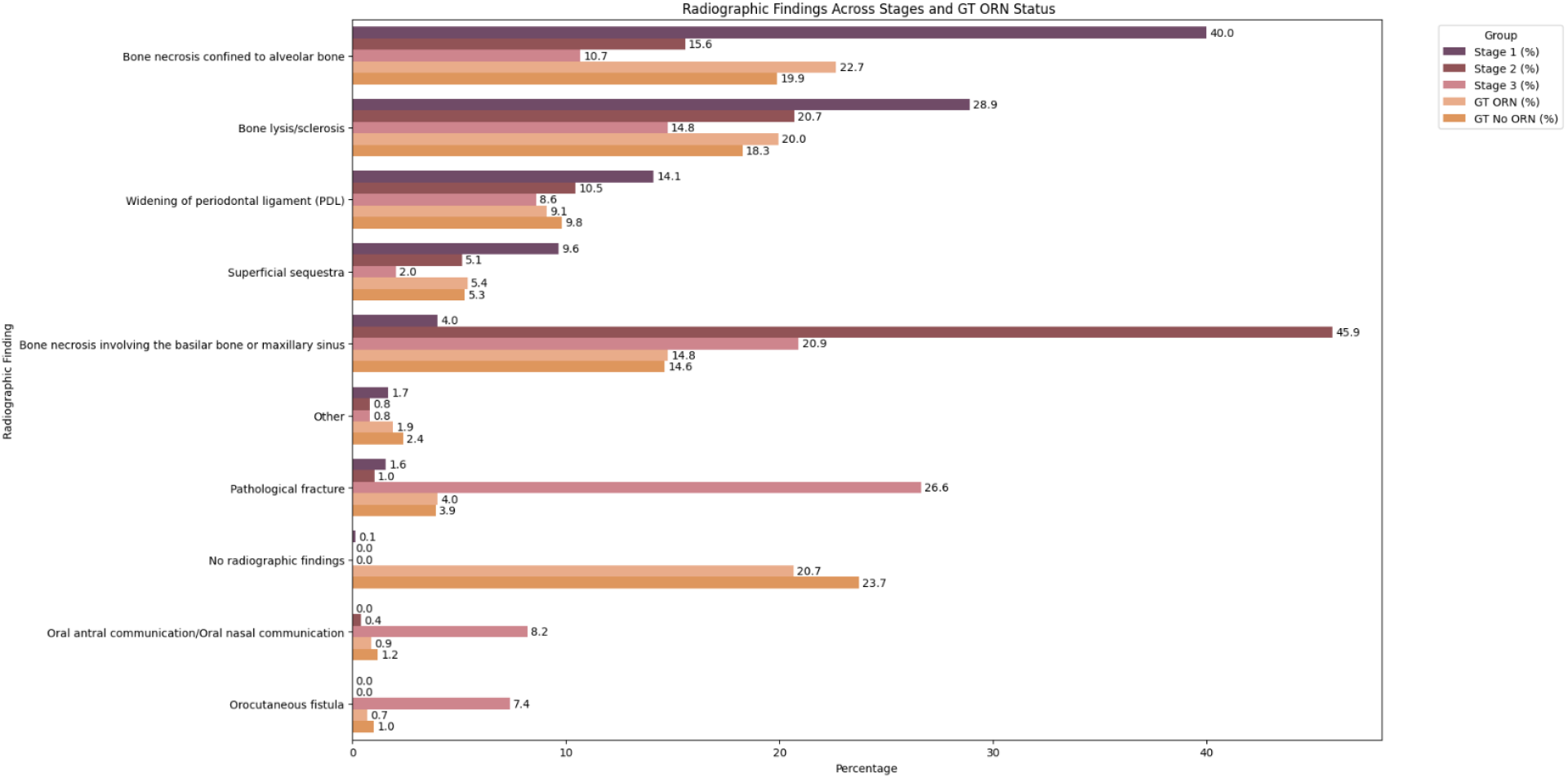
Distribution of radiographic features selected by ORNJ stage as determined by survey participants using CT and OPG images.

### 3.5 Inter and intra-rater variability

Slight inter-rater agreement in detecting ORNJ (i.e., ORNJ vs. non-ORN) was observed with median (IQR) Fleiss kappa values of 0.38 (0.22) and 0.21 (0.29) for the entire set, the non-ORNJ cases subset and the ORNJ cases subset, respectively. Slight to fair inter-rater agreement in severity classification of ORNJ (i.e., stage 0/1 vs. stage 2 vs. stage 3) was observed with median (IQR) Fleiss kappa values of 0.22, 0.13, and 0.05 for stages 0/1, 2, and 3, respectively. Slight intra-rater agreement for physician detection of ORNJ was observed, with a median (IQR) Cohens’ kappa value of 0.08 (0.17).

## 4. Discussion

This study reveals significant variability in physician-based diagnostic accuracy for ORNJ detection and severity classification across clinical specialties, emphasizing the need for a standardized approach. Residents, radiation oncologists, and neuroradiologists displayed notable differences in agreement and accuracy, highlighting gaps in diagnostic consistency, which could potentially be addressed with further training. Despite these differences, none of the individual specialties demonstrated statistically significant diagnostic ability beyond chance after Bonferroni correction (p > 0.05 for all). Even among the highest-performing groups, such as radiation oncologists (AUC = 0.552, p = 0.062) and dentists (AUC = 0.549, p = 0.133), performance did not significantly exceed random classification. This reinforces the inherent difficulty of ORN detection using radiographic imaging alone, underscoring the need for clinical correlation and additional diagnostic tools. However, when CT and OPG imaging were combined, all specialties demonstrated significantly improved diagnostic performance (p < 0.001 for all), with AUC values exceeding chance levels (AUC range: 0.786–0.977). Surgeons exhibited the highest AUC (0.977), while residents had the lowest (0.786), suggesting that multimodal imaging benefits all groups, regardless of specialty. Notably, DeLong’s test revealed no statistically significant differences between specialties (p > 0.05 for all), indicating that paired imaging improved diagnostic accuracy across all groups to a similar extent. These findings highlight the diagnostic advantage of multimodal imaging for ORN detection and suggest that systematic integration of CT and OPG could enhance clinical decision-making across specialties, enabling earlier diagnosis and better patient outcomes.

These trends likely reflect differences in training focus, clinical exposure, and imaging interpretation experience across specialties. Dentists and surgeons, who demonstrated the highest accuracy with OPG imaging, routinely assess dental radiographs and are accustomed to identifying subtle osseous changes in the maxillofacial region. Conversely, residents, despite their broad medical training, may lack extensive experience in ORNJ-specific imaging, leading to higher sensitivity but lower specificity due to a tendency to err on the side of caution. Radiation oncologists, while regularly managing patients at risk for ORNJ, often rely on clinical rather than radiographic assessment for disease progression, potentially explaining their moderate diagnostic performance. Neuroradiologists, who typically interpret a wide range of head and neck imaging, may face challenges in distinguishing ORNJ from other conditions, such as osteomyelitis or malignancy, contributing to their lower specificity. General practitioners, with the lowest performance overall, likely have the least exposure to ORNJ cases and limited specialized radiographic training, leading to higher misclassification rates. This underscores the need for targeted training programs focused on ORNJ-specific radiographic features and the development of a standardized diagnostic framework that incorporates guidelines for multimodal imaging interpretation, decision-support tools, and interdisciplinary collaboration to enhance consistency across specialties.

Our findings align with previous studies that have highlighted the challenges of diagnosing ORNJ due to its subtle radiographic features.^20^ For instance, a study by Yilmaz et al. discusses the difficulty in distinguishing ORNJ from other conditions because of overlapping clinical and radiological characteristics.^21^ Additionally, many earlier works have focused on single imaging modalities, such as computed tomography (CT) or orthopantomography (OPG), without rigorous comparison across multiple specialties. For example, a study by Miyamoto et al. evaluated the diagnostic aspects of ORN using several imaging modalities but did not extensively compare diagnostic performance across different medical specialties.^22^

Distinguishing ORN from recurrent malignancy remains a critical challenge in post-treatment head and neck cancer imaging. Cheng et al. emphasize the role of hybrid imaging techniques, such as PET/CT, in differentiating post-radiation bone necrosis from tumor recurrence.^23^ Similarly, Alhilali et al. discuss the limitations of conventional MRI and CT in distinguishing ORN from recurrent disease, given overlapping radiographic features such as cortical bone erosion and marrow signal changes.^24^ However, this study did not aim to assess the performance of imaging in differentiating ORN from recurrence, since all the ORN cases were clinically confirmed. Instead, our focus was on evaluating physician interpretation of radiographic findings specifically for ORN detection, without assessing neoplastic progression.

While no prior studies have comprehensively benchmarked ORN detection across multiple specialties, previous research on medication-related osteonecrosis of the jaw (MRONJ) offers insights into the radiographic characteristics of bone necrosis. MRONJ and ORN share similar radiographic hallmarks, including osteosclerosis, cortical disruption, and pathological fractures. However, MRONJ tends to present with more prominent sequestration and periosteal reactions due to its distinct pathophysiology, involving bisphosphonates or other antiresorptive agents. These differences underscore the need for specialized imaging-based criteria tailored to radiation-induced versus medication-induced necrosis. Future research could explore whether radiomics-based methods used in MRONJ detection are applicable to ORN, particularly in differentiating early-stage disease from recurrence.

As shown in previous studies^10,12,23^, image-based information is fundamental in the detection of mandibular bone damage. Leveraging Watson et al.^7^, we found that incorporating radiographic findings into ORNJ severity classification improved diagnostic performance. Unlike studies relying solely on clinical assessments our results underscore the value of multiple imaging modalities in reducing diagnostic variability.^25^ This supports the conclusions of Wang et al., who reported that multimodal 2D/3D imaging approaches can improve diagnostic precision and facilitate optimal clinical outcomes.^26^

Several limitations may be noted. Specialty representation was variable in terms of observer number, which could have introduced bias in the observed variability of diagnostic performance.^27^ This may have limited the generalizability of our findings across specialties, as certain performance-driving expertise might not have been fully captured in the analyses, including variables beyond ORNJ case volume and years of experience—such as differences in institutional training, prior exposure to ORNJ imaging, reliance on adjunct imaging modalities, and cognitive heuristics in radiographic interpretation. Additionally, the uneven distribution of specialties could have skewed the comparative accuracy rates, potentially overstating or understating the performance of specific specialties. Where suitable, percentages rather than response counts were considered to address this limitation. Second, despite the onboarding training session, the fact that the paired modality assessments were presented at the end (i.e., after the single modality assessments) could have led to additional unintended training of participants, thus potentially contributing to the significantly better ORNJ discrimination results obtained with paired images. Third, reviewer fatigue over the course of the study, particularly during prolonged review sessions, may have influenced accuracy rates and contributed to diagnostic variability. However, to mitigate fatigue-related bias, this study incorporated randomized image ordering, ensuring that case order did not systematically influence diagnostic performance. While this approach helped reduce sequential bias, attention fatigue may still have played a role in the variability observed across specialties. Fourth, this study utilized a classification system, ClinRad, that requires both clinical and radiographic findings to arrive at a definitive stage. Indeed, we observed more variability in diagnosing stage 0/1 ORN than more advanced stages, which makes logical sense, as stage 2 and 3 represent more advanced stages with greater bone destruction that is quite evident radiographically. In the earlier stages of ORN, radiographic findings are less pronounced or even absent, with clinical findings and background more relevant to arrive at a diagnosis of ORN than in later stages. Finally, although the review sessions were initially planned in-person to ensure equal external conditions (e.g., lighting) across all participants, several physician observers required asynchronous review, affording differential environmental conditions.

Despite these caveats, the clinical implications of this study are significant. Variability in ORNJ detection across specialties highlights the critical need for interdisciplinary collaboration and standardized training. Paired imaging modalities have proven effective but require clinicians to interpret diverse radiographic data proficiently. By adopting standardized imaging protocols and promoting consistent training, clinical practice can evolve to reduce diagnostic disparities and improve outcomes for patients with ORN.

To our knowledge, this effort represents the 1^st^ reported prospective radiographic assessment of diagnostic performance for image-detected ORN. As “imaging only” mandibular degradation is now deemed by ASCO/ISOO/MASCC guidance as a diagnostic criteria for ORNJ^28^ and consensus guidance recommends image-based staging,^29^ our data provides the first extant and definitive initial benchmarking, and, importantly, suggests that matched 2D (OPG) and 3D (CT) bone assessment leads to improved detection and monitoring capability of ORN. At the same time, our findings emphasize that ORN can exist without radiographic findings, making early-stage detection more challenging without the inclusion of clinical findings. We observed increased variability in diagnosing stage 0/1 ORN compared to more advanced stages, further supporting the notion that radiographic findings alone may be insufficient for early ORN diagnosis. Future studies will incorporate clinical findings alongside radiographic assessments to enhance diagnostic accuracy and reliability.

In conclusion, this study provides a critical benchmark for clinician performance in ORNJ detection and severity classification using the ClinRad system. By identifying variability across specialties and demonstrating the advantages of paired imaging modalities, we lay the groundwork for standardizing diagnostic practices and exploring innovative approaches to enhance accuracy and consistency.

## Data Availability

Data produced are available online at: 10.6084/m9.figshare.28395047. Any additional data produced in the present study are available upon reasonable request to the authors.

## References

1. Barsouk A, Aluru JS, Rawla P, Saginala K, Barsouk A. Epidemiology, Risk Factors, and Prevention of Head and Neck Squamous Cell Carcinoma. Medical Sciences. 2023;11(2):42. doi:10.3390/medsci11020042

2. Faraji F, Rettig EM, Tsai HL, et al. The prevalence of human papillomavirus in oropharyngeal cancer is increasing regardless of sex or race, and the influence of sex and race on survival is modified by human papillomavirus tumor status. Cancer. 2019;125(5):761–769. doi:10.1002/cncr.31841

3. Senkomago V, Henley SJ, Thomas CC, Mix JM, Markowitz LE, Saraiya M. Human Papillomavirus-Attributable Cancers - United States, 2012-2016. MMWR Morb Mortal Wkly Rep. 2019;68(33):724–728. doi:10.15585/mmwr.mm6833a3

4. Frankart AJ, Frankart MJ, Cervenka B, Tang AL, Krishnan DG, Takiar V. Osteoradionecrosis: Exposing the Evidence Not the Bone. International Journal of Radiation Oncology Biology Physics. 2021;109(5):1206–1218. doi:10.1016/j.ijrobp.2020.12.043

5. Patel V, Ormondroyd L, Lyons A, McGurk M. The financial burden for the surgical management of osteoradionecrosis. British Dental Journal. 2017;222(3):177–180. doi:10.1038/sj.bdj.2017.121

6. O’Dell K, Sinha U. Osteoradionecrosis. Oral Maxillofac Surg Clin North Am. 2011;23(3):455–464. doi:10.1016/j.coms.2011.04.011

7. Watson EE, Hueniken K, Lee J, et al. Development and Standardization of an Osteoradionecrosis Classification System in Head and Neck Cancer: Implementation of a Risk-Based Model. J Clin Oncol. 2024;42(16):1922–1933. doi:10.1200/JCO.23.01951

8. Peterson DE, Koyfman SA, Yarom N, et al. Prevention and Management of Osteoradionecrosis in Patients With Head and Neck Cancer Treated With Radiation Therapy: ISOO-MASCC-ASCO Guideline. JCO. 2024;42(16):1975–1996. doi:10.1200/JCO.23.02750

9. Consortium TIO. International Expert-Based Consensus Definition, Staging Criteria, and Minimum Data Elements for Osteoradionecrosis of the Jaw: An Inter-Disciplinary Modified Delphi Study. medRxiv. Published online April 9, 2024:2024.04.07.24305400. doi:10.1101/2024.04.07.24305400

10. Joint Head and Neck Radiation Therapy-MRI Development Cooperative, Mohamed ASR, He R, et al. Quantitative Dynamic Contrast-Enhanced MRI Identifies Radiation-Induced Vascular Damage in Patients With Advanced Osteoradionecrosis: Results of a Prospective Study. Int J Radiat Oncol Biol Phys. 2020;108(5):1319–1328. doi:10.1016/j.ijrobp.2020.07.029

11. Cooperative JH and NMRD Dijk LV van, Ventura J, et al. Black bone MRI morphometry for mandibular cortical bone measurement in head and neck cancer patients: Prospective method comparison with CT. Published online July 26, 2020. doi:10.1101/2020.07.21.20154880

12. Elhalawani H, Volpe S, Barua S, et al. Exploration of an Early Imaging Biomarker of Osteoradionecrosis in Oropharyngeal Cancer Patients: Case-Control Study of the Temporal Changes of Mandibular Radiomics Features. International Journal of Radiation Oncology, Biology, Physics. 2018;100(5):1363–1364. doi:10.1016/j.ijrobp.2017.12.146

13. Barua S, Elhalawani H, Volpe S, et al. Computed Tomography Radiomics Kinetics as Early Imaging Correlates of Osteoradionecrosis in Oropharyngeal Cancer Patients. Front Artif Intell. 2021;4. doi:10.3389/frai.2021.618469

14. Sharma A, Minh Duc NT, Luu Lam Thang T, et al. A Consensus-Based Checklist for Reporting of Survey Studies (CROSS). J Gen Intern Med. 2021;36(10):3179–3187. doi:10.1007/s11606-021-06737-1

15. STARD for Abstracts: essential items for reporting diagnostic accuracy studies in journal or conference abstracts | EQUATOR Network. Accessed February 4, 2025. https://www.equator-network.org/reporting-guidelines/stard-abstracts/

16. G S. Sample size requirements for the design of reliability studies: precision consideration. PubMed. Accessed November 22, 2024. https://pubmed.ncbi.nlm.nih.gov/24338600/

17. A D, M E. A goodness-of-fit approach to inference procedures for the kappa statistic: confidence interval construction, significance-testing and sample size estimation. PubMed. Accessed November 22, 2024. https://pubmed.ncbi.nlm.nih.gov/1410963/

18. Landis JR, Koch GG. An Application of Hierarchical Kappa-type Statistics in the Assessment of Majority Agreement among Multiple Observers. Biometrics. 1977;33(2):363–374. doi:10.2307/2529786

19. Sim J, Wright CC. The kappa statistic in reliability studies: use, interpretation, and sample size requirements. Physical therapy. 2005;85(3):257–268.

20. Busra Yilmaz DDS, Efsun Somay DDS, Ahmet Kucuk MD, Berrin Pehlivan MD, Ugur Selek MD, Erkan Topkan MD. Challenges in the Radiological Diagnosis of Osteoradionecrosis of the Jaw in Head and Neck Cancer Patients. Exon Publications. Published online October 28, 2022:1-22. doi:10.36255/osteoradionecrosis-radiological-diagnosis

21. B Y, E S, A K, B P, U S, E T. Challenges in the Radiological Diagnosis of Osteoradionecrosis of the Jaw in Head and Neck Cancer Patients. PubMed. Accessed February 6, 2025. https://pubmed.ncbi.nlm.nih.gov/37756428/

22. Miyamoto I, Tanaka R, Kogi S, et al. Clinical Diagnostic Imaging Study of Osteoradionecrosis of the Jaw: A Retrospective Study. J Clin Med. 2021;10(20):4704. doi:10.3390/jcm10204704

23. Cheng NM, Lin CY, Liao CT, Tsan DL, Ng SH, Yen TC. The added values of 18F-FDG PET/CT in differentiating cancer recurrence and osteoradionecrosis of mandible in patients with treated oral squamous cell carcinoma. EJNMMI Research. 2023;13(1):25. doi:10.1186/s13550-023-00965-8

24. Alhilali L, Reynolds AR, Fakhran S. Osteoradionecrosis after Radiation Therapy for Head and Neck Cancer: Differentiation from Recurrent Disease with CT and PET/CT Imaging. Published online July 1, 2014. doi:10.3174/ajnr.A3879

25. Osteoradionecrosis: Exposing the Evidence Not the Bone - International Journal of Radiation Oncology, Biology, Physics. Accessed December 19, 2024. https://www.redjournal.org/article/S0360-3016%2820%2934739-8/fulltext

26. Wang Y, Turkstani H, Alfaifi A, Akintoye SO. Diagnostic and Therapeutic Approaches to Jaw Osteoradionecrosis. Diagnostics. 2024;14(23):2676. doi:10.3390/diagnostics14232676

27. Frankart AJ, Frankart MJ, Cervenka B, Tang AL, Krishnan DG, Takiar V. Osteoradionecrosis: Exposing the Evidence Not the Bone. Accessed December 19, 2024.https://www.redjournal.org/article/S0360-3016%2820%2934739-8/fulltext?

28. Prevention and Management of Osteoradionecrosis in Patients With Head and Neck Cancer Treated With Radiation Therapy: ISOO-MASCC-ASCO Guideline | Journal of Clinical Oncology. Accessed February 6, 2025. https://ascopubs.org/doi/full/10.1200/JCO.23.02750

29. Moreno AC, Watson EE, Humbert-Vidan L, et al. International Expert-Based Consensus Definition, Classification Criteria, and Minimum Data Elements for Osteoradionecrosis of the Jaw: An Inter-Disciplinary Modified Delphi Study. International Journal of Radiation Oncology*Biology*Physics. Published online January 16, 2025. doi:10.1016/j.ijrobp.2024.12.017

