## Supplemental Material for "Radiographic classification of mandibular osteoradionecrosis: A blinded prospective multi-disciplinary interobserver diagnostic performance study"

**Supplementary Material – Table of Contents**


**Supplement A. Checklist for Reporting of Survey Studies (CROSS) and Abstracts (STARD)**

**Supplement B. Survey participants**

**Supplement C. Inclusion/exclusion subject criteria flowchart**

**Supplement D. Patient cohort characteristics**

**Supplement E. ClinRad ORN stages used in this Survey**

**Supplement F. CT acquisition parameters**

**Supplement G. Qualtrics Survey presented to participants.**

**Supplement H. ROC analyses results.**

**Supplement I. DeLong's Test results for ROC Comparison**

**Supplement J. Confidence in ORN staging analyses.**

### Supplement A. Checklist for Reporting of Survey Studies (CROSS) and Abstracts (STARD)

Table A1. Checklist for Reporting of Survey Studies (CROSS)

| Section/topic | Item | Item description | Reported on page # |
|---|---|---|---|
| **Title and abstract** | | | |
| Title and abstract | 1a | State the word "survey" along with a commonly used term in title or abstract to introduce the study's design. | 1 |
| | 1b | Provide an informative summary in the abstract, covering background, objectives, methods, findings/results, interpretation/discussion, and conclusions. | 1 |
| **Introduction** | | | |
| Background | 2 | Provide a background about the rationale of study, what has been previously done, and why this survey is needed. | 1-2 |
| Purpose/aim | 3 | Identify specific purposes, aims, goals, or objectives of the study. | 2 |
| **Methods** | | | |
| Study design | 4 | Specify the study design in the methods section with a commonly used term (e.g., cross-sectional or longitudinal). | 2 |
| | 5a | Describe the questionnaire (e.g., number of sections, number of questions, number and names of instruments used). | 2/3 |
| Data collection methods | 5b | Describe all questionnaire instruments that were used in the survey to measure particular concepts. Report target population, reported validity and reliability information, scoring/classification procedure, and reference links (if any). | 2/3 |
| | 5c | Provide information on pretesting of the questionnaire, if performed (in the article or in an online supplement). Report the method of pretesting, number of times questionnaire was pre-tested, number and demographics of participants used for pretesting, and the level of similarity of demographics between pre-testing participants and sample population. | N/A |
| | 5d | Questionnaire, if possible, should be fully provided (in the article, or as appendices or as an online supplement). | Supplement D |
| Sample characteristics | 6a | Describe the study population (i.e., background, locations, eligibility criteria for participant inclusion in survey, exclusion criteria). | 2 |
| | 6b | Describe the sampling techniques used (e.g., single stage or multistage sampling, simple random sampling, stratified sampling, cluster sampling, convenience sampling). Specify the locations of sample participants whenever clustered sampling was applied. | 2 |
| | 6c | Provide information on sample size, along with details of sample size calculation. | 6 |
| | 6d | Describe how representative the sample is of the study population (or target population if possible), particularly for population-based surveys. | 4 |
| Survey administration | 7a | Provide information on modes of questionnaire administration, including the type and number of contacts, the location where the survey was conducted (e.g., outpatient room or by use of online tools, such as SurveyMonkey). | 2 |
| | 7b | Provide information of survey's time frame, such as periods of recruitment, exposure, and follow-up days. | 3 |
| | 7c | Provide information on the entry process: | 2/3 |

| | | | |
|---|---|---|---|
| | | –>For non-web-based surveys, provide approaches to minimize human error in data entry.<br>–>For web-based surveys, provide approaches to prevent "multiple participation" of participants. | |
| Study preparation | 8 | Describe any preparation process before conducting the survey (e.g., interviewers' training process, advertising the survey). | 3 |
| Ethical considerations | 9a | Provide information on ethical approval for the survey if obtained, including informed consent, institutional review board [IRB] approval, Helsinki declaration, and good clinical practice [GCP] declaration (as appropriate). | 2 |
| | 9b | Provide information about survey anonymity and confidentiality and describe what mechanisms were used to protect unauthorized access. | 3 |
| Statistical analysis | 10a | Describe statistical methods and analytical approach. Report the statistical software that was used for data analysis. | 3 |
| | 10b | Report any modification of variables used in the analysis, along with reference (if available). | 3/4 |
| | 10c | Report details about how missing data was handled. Include rate of missing items, missing data mechanism (i.e., missing completely at random [MCAR], missing at random [MAR] or missing not at random [MNAR]) and methods used to deal with missing data (e.g., multiple imputation). | N/A |
| | 10d | State how non-response error was addressed. | 12 |
| | 10e | For longitudinal surveys, state how loss to follow-up was addressed. | N/A |
| | 10f | Indicate whether any methods such as weighting of items or propensity scores have been used to adjust for non-representativeness of the sample. | N/A |
| | 10g | Describe any sensitivity analysis conducted. | 3 |
| **Results** | | | |
| Respondent characteristics | 11a | Report numbers of individuals at each stage of the study. Consider using a flow diagram, if possible. | 3 |
| | 11b | Provide reasons for non-participation at each stage, if possible. | N/A |
| | 11c | Report response rate, present the definition of response rate or the formula used to calculate response rate. | Supplement B |
| | 11d | Provide information to define how unique visitors are determined. Report number of unique visitors along with relevant proportions (e.g., view proportion, participation proportion, completion proportion). | 6 |
| Descriptive results | 12 | Provide characteristics of study participants, as well as information on potential confounders and assessed outcomes. | Supplement B |
| Main findings | 13a | Give unadjusted estimates and, if applicable, confounder-adjusted estimates along with 95% confidence intervals and p-values. | N/A |
| | 13b | For multivariable analysis, provide information on the model building process, model fit statistics, and model assumptions (as appropriate). | N/A |
| | 13c | Provide details about any sensitivity analysis performed. If there are considerable amount of missing data, report sensitivity analyses comparing the results of complete cases with that of the imputed dataset (if possible). | Supplement E |
| **Discussion** | | | |

| | | | |
|---|---|---|---|
| Limitations | 14 | Discuss the limitations of the study, considering sources of potential biases and imprecisions, such as non-representativeness of sample, study design, important uncontrolled confounders. | 9 |
| Interpretations | 15 | Give a cautious overall interpretation of results, based on potential biases and imprecisions and suggest areas for future research. | 10 |
| Generalizability | 16 | Discuss the external validity of the results. | 10 |
| **Other sections** | | | |
| Role of funding source | 17 | State whether any funding organization has had any roles in the survey's design, implementation, and analysis. | 1 |
| Conflict of interest | 18 | Declare any potential conflict of interest. | 1 |
| Acknowledgements | 19 | Provide names of organizations/persons that are acknowledged along with their contribution to the research. | 1 |

Table A2. STARD checklist for abstracts: essential items for reporting diagnostic accuracy studies in journal or conference abstracts

| Section | Item |
|---|---|
| | Identification as a study of diagnostic accuracy using at least one measure of accuracy (such as sensitivity, specificity, predictive values, or AUC) |
| **Background and Objectives** | Study objectives – pg 1 and 3 |
| **Methods** | Data collection: whether this was a prospective or **retrospective** study – pg 1 and 3 |
| | Eligibility criteria for participants and settings where the data were collected – pg 4 |
| | Whether participants formed a consecutive, random, or convenience series – pg 5 |
| | Description of the index test and reference standard - pg 5 |
| **Results** | Number of participants with and without the target condition included in the analysis – pg 7 |
| | Estimates of diagnostic accuracy and their precision (such as 95% confidence intervals) - pg 7 and 8 |
| **Discussion** | General interpretation of the results – pg 12 |
| | Implications for practice, including the intended use of the index test – pg 13 |
| **Registration** | Registration number and name of registry – n/a |

**Supplement B. Survey participants**

Table B1. Familiarity of survey participant with staging system and software used.

Table B1a: ORN staging approach per specialty.

| ORN Staging Approach | Uses a formal staging system | Does not use staging system | Uses ClinRad staging system |
|---|---|---|---|
| Radiation Oncologist | 75% | 25% | 50% |
| Dentist | 33.30% | 66.66% | 0% |
| Surgeon | 50% | 50% | 50% |
| Neuroradiologist | 0% | 100% | 0% |
| Resident | 0% | 100% | 0% |

Table B1b: Percent of participants who use ClinRad staging system vs others or none.

| Percent of Doctors who use ClinRad as a staging system | Percent of Doctors who use Marx staging system | Percent of Doctors who use Tsai staging system | Percent of Doctors who used CTCAE v5.0 staging system | Percent of Doctors who do NOT use a staging system |
|---|---|---|---|---|
| 14.30% | 4.76% | 4.76% | 4.76% | 66.66% |

Table B1c: Percent of participants familiarity with RayStation.

| Familiar with Ray Station | Percent |
|---|---|
| Very familiar | 38.10% |
| Somewhat familiar | 9.52% |
| Neutral | 4.76% |
| Not too familiar | 14.28% |
| Not familiar at all | 33.33% |

**Supplement C. Inclusion/exclusion subject criteria flowchart**

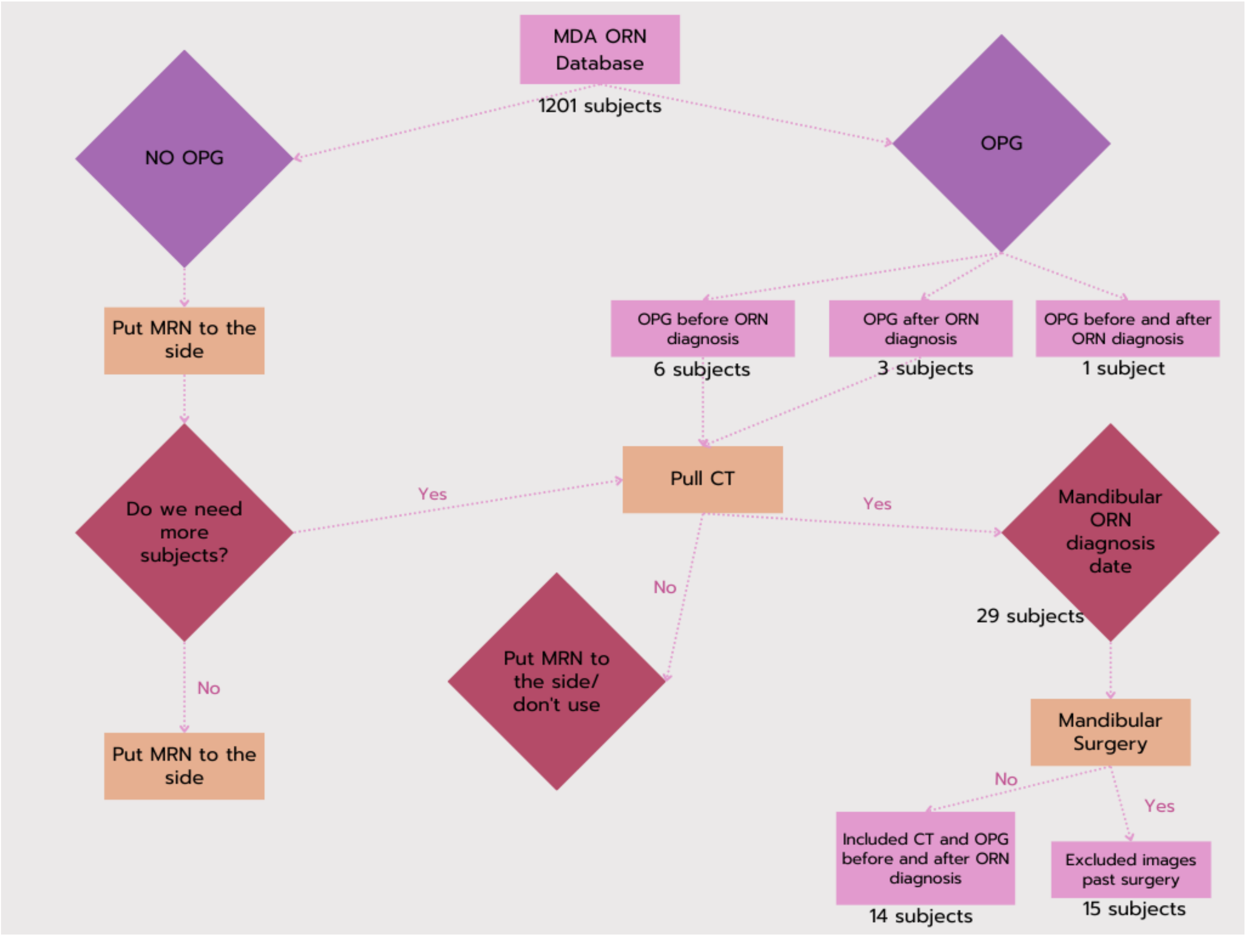


Figure C1. Flowchart depicting the inclusion and exclusion criteria for subject selection based on the availability of OPG and CT imaging in the MDA ORN database.

### Supplement D. Patient cohort characteristics

Table D1. Patient clinical and demographic characteristics.

| Category | N (%) |
|---|---|
| Total | 30 |
| Age (median, range) | 67 (59-72) |
| Sex | |
| *Male / Female* | 28 (43.1) / 2 (3.1) |
| Dental Extractions | |
| *Yes / No* | 19 (29.2%) / 8 (12.3%) |
| HPV Status | |
| *Positive / Negative* | 10 (15.4) / 5 (7.7) |
| Medical History | |
| *Former smoker* | 10 (15.4) |
| *Alcohol use* | 10 (15.4) |
| *Drug use* | 5 (7.7) |

### Supplement E. ClinRad ORN stages used in this Survey

Table E1. Description of the ClinRad[7] ORN stages used in this study.

| ClinRad Scale | Scale Used for Study | Description |
|---|---|---|
| Minor Bone Spicules (Pre-ORN Stage) | Not used | Distinct from ORN, occurs with superficial mobile spicules or sequestra within mucosa, no radiographic evidence of bone necrosis. |
| Stage 0 | Used | Radiographic evidence of bone necrosis confined to alveolar bone, with features like bone lysis/sclerosis, intact mucosa. |
| Stage 1 | Not used | Clinical signs of ORN with exposed bone or intact mucosa, confined to alveolar bone. May need minor surgical or medical intervention. |
| Stage 2 | Used | Radiographic evidence involving basilar bone or maxillary sinus, with exposed bone or intact mucosa. Intermediate surgical intervention may be needed. |
| Stage 3 | Used | Advanced ORN with pathologic fractures, fistulas, or communication issues. Requires reconstructive surgical intervention. |

| Stage | Radiographic Findings | Example Image |
|---|---|---|
| Stage 0 | - **Bone necrosis confined to ALVEOLAR BONE**<br>- Bone lysis/sclerosis<br>- Widening periodontal ligament (PDL) space<br>- Absence of osseous filling of extraction sockets | 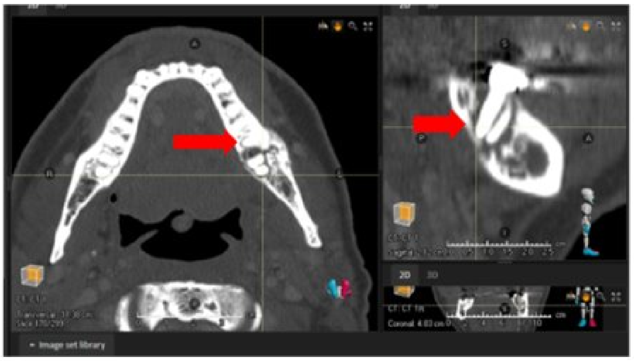 |
| Stage 1 | - None or as stage 0 | 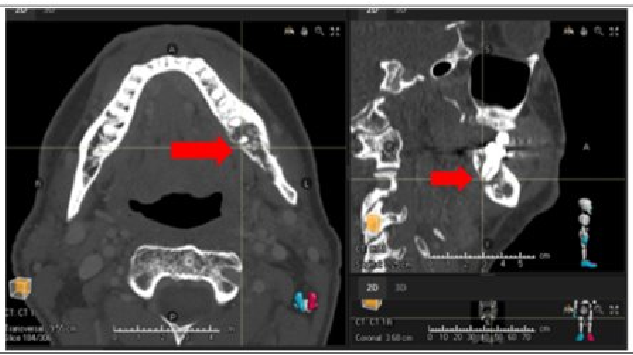 |
| Stage 2 | - Bone necrosis involving **BASILAR BONE or MAXILLARY SINUS** | 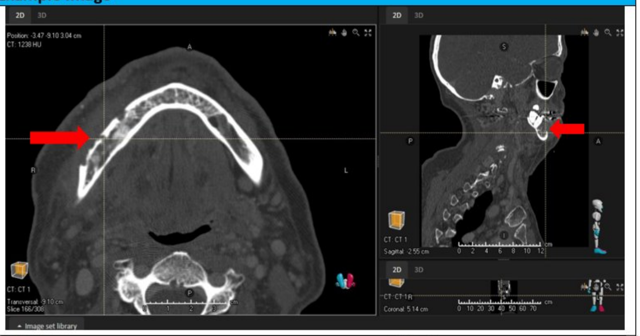 |
| Stage 3 | - One or more of the following,<br>- Pathological fracture<br>- Orcocutaneous fistula<br>- Oral antral communication/oral nasal communication | 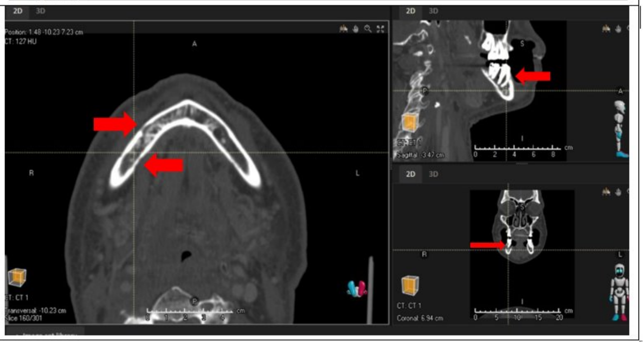 |

Figure E1. Examples shown to survey participants during pre-survey education session to demonstrate the different stages of the ClinRad ORN classification system.

**Supplement F. CT acquisition parameters**

| Parameter | **Median** | **IQR** | **Range** |
|---|---|---|---|
| KVP | 120 | (120.0, 120.0) | (120.0, 120.0) |
| Slice Thickness (mm) | 1.25 | (1.25, 1.25) | (0.625, 1.5) |
| Exposure Time (ms) | 1000 | (1000.0, 1000.0) | (1000, 1825) |
| X-Ray Tube Current (mA) | 313 | (259.0, 393.0) | (159, 408) |
| Rows | 512 | (512.0, 512.0) | (512, 512) |
| Columns | 512 | (512.0, 512.0) | (512, 512) |
| Pixel Spacing (mm) | 0.488 | (0.488, 0.488) | (0.488, 0.488) |
| Reconstruction Diameter (mm) | 250 | (250.0, 250.0) | (250.0, 270.0) |
| Axial Slices | 297 | (279.0, 312.0) | (48, 468) |

**Supplement G. Qualtrics Survey presented to participants.**

Study participants completed a Qualtrics survey consisting of three sections: a pre-survey (Table G1), a main survey (Table G2), and a paired survey (Table G3). The main survey was designed to capture their diagnostic and staging decisions, the decision-making process (including identification of observed anatomical abnormalities) and confidence levels in each assessment using a Likert scale. The paired survey assessed diagnostic consistency using combined imaging modalities (OPG and CT), focusing on abnormalities, ORN status, staging, and confidence.

Table G1. Pre-survey

| **Question Number** | **Question** | **Response Type** | **Response Options** |
|---|---|---|---|
| 1 | Reader's Unique Identifier | Open-ended | N/A |
| 2 | How would you define ORN? | Open-ended | N/A |
| 3 | How many ORN cases do you see annually? | Single choice (Multiple choice) | None<br>1-5<br>5-10<br>10-20<br>20+ |
| 4 | How do you typically stage ORN? List staging systems if applicable. | Open-ended | N/A |
| 5 | How familiar are you with RayStaytion? | Single choice (Multiple choice) | Very familiar<br>Somewhat familiar<br>Neutral<br>Not too familiar<br>Not familiar at all |
| 6 | How familiar are you with RayStaytion? | Open-ended | N/A |
| 7 | Years of experience in specialty: | Single choice (multiple choice) | Resident<br>1-5<br>5-10 |

| | | | |
|---|---|---|---|
| | | | 10-15<br>15-20<br>20+ |

Table G2. Main survey

| Question Number | Question | Response Type | Response Options |
|---|---|---|---|
| 1 | Reader's Unique Identifier | Open-ended | N/A |
| 2 | Please enter subject ID | Open-ended | N/A |
| 3 | Modality used for radiographic evaluation | Single choice (Multiple choice) | Orthopantomogram (OPG)<br>CT head and/or neck with contrast<br>CT head and/or neck without contrast |
| 4 | Are there any abnormalities present? | Yes/No | Yes<br>No |
| 5 | Select the most accurate statements that describe the radiographic findings. | Choose multiple answers if applicable | Superficial sequestra<br>Bone necrosis confined to alveolar bone<br>Bone necrosis involving the basilar bone or maxillary sinus<br>Bone lysis/sclerosis<br>Widening of periodontal ligament (PDL)<br>Pathological fracture<br>Orocutaneous fistula<br>Oral antral communication/Oral nasal communication fracture<br>*No radiographic findings<br>Other (option to write response in) |
| 6 | How confident are you that there are abnormalities present? | Likert scale (Levels of confidence) | 5- Not at all confident<br>5- Slightly confident<br>5- Somewhat confident<br>5- Fairly confident<br>5- Completely confident |
| 7 | Does the subject have ORN? | Yes/No | Yes<br>Maybe<br>No |
| 8 | How confident are you that the subject does/does not have ORN? | Likert scale (Levels of confidence) | 5- Not at all confident<br>5- Slightly confident<br>5- Somewhat confident<br>5- Fairly confident |

| | | | |
|---|---|---|---|
| | | | 5- Completely confident |
| 9 | Using the RadORN (Watson Scale) please stage the level of ORN. | Single choice (Multiple choice) | No ORN<br>Stage 0/1<br>Stage 2<br>Stage 3 |
| 10 | How confident are you with this staging? | Likert scale (Levels of confidence) | 5- Not at all confident<br>5- Slightly confident<br>5- Somewhat confident<br>5- Fairly confident<br>5- Completely confident |

Table G3. Paired survey

| **Question Number** | **Question** | **Response Type** | **Response Options** |
|---|---|---|---|
| 1 | Reader's Unique Identifier | Open-ended | N/A |
| 2 | Please enter subject ID (OPG) | Open-ended | N/A |
| 3 | Modality used for radiographic evaluation | Single choice (Multiple choice) | Orthopantomogram (OPG)<br>CT head and/or neck with contrast<br>CT head and/or neck without contrast |
| 4 | Are there any abnormalities present? | Yes/No | Yes<br>No |
| 5 | Select the most accurate statements that describe the radiographic findings. | Choose multiple answers if applicable | Superficial sequestra<br>Bone necrosis confined to alveolar bone<br>Bone necrosis involving the basilar bone or maxillary sinus<br>Bone lysis/sclerosis<br>Widening of periodontal ligament (PDL)<br>Pathological fracture<br>Orocutaneous fistula<br>Oral antral communication/Oral nasal communication fracture<br>*No radiographic findings<br>Other (option to write response in) |
| 6 | How confident are you that there are abnormalities present? | Likert scale (Levels of confidence) | 5- Not at all confident<br>5- Slightly confident<br>5- Somewhat confident |

| | | | |
|---|---|---|---|
| | | | 5- Fairly confident<br>5- Completely confident |
| 7 | Does the subject have ORN? | Yes/No | Yes<br>Maybe<br>No |
| 8 | How confident are you that the subject does/does not have ORN? | Likert scale (Levels of confidence) | 5- Not at all confident<br>5- Slightly confident<br>5- Somewhat confident<br>5- Fairly confident<br>5- Completely confident |
| 9 | Using the RadORN (Watson Scale) please stage the level of ORN. | Single choice (Multiple choice) | No ORN<br>Stage 0/1<br>Stage 2<br>Stage 3 |
| 10 | How confident are you with this staging? | Likert scale (Levels of confidence) | 5- Not at all confident<br>5- Slightly confident<br>5- Somewhat confident<br>5- Fairly confident<br>5- Completely confident |

### Supplement H. ROC analyses results

Table H1. ORN detection performance results by specialty and years of experience of survey participants when using CT images only.

| **Specialty (*years of experience*)** | **TP** | **FP** | **TN** | **FN** | **Accuracy (mean, SD)** | **Sensitivity** | **Specificity** | **Precision** |
|---|---|---|---|---|---|---|---|---|
| Dentist | | | | | | | | |
| *5-10* | 13 | 13 | 19 | 18 | 0.51 (±0.07) | 0.42 | 0.59 | 0.50 |
| *10-15* | 24 | 17 | 14 | 7 | 0.61 (±0.13) | 0.77 | 0.45 | 0.58 |
| *15-20* | 19 | 21 | 9 | 14 | 0.44 (±0.11) | 0.57 | 0.30 | 0.47 |
| Radiation Oncologist | | | | | | | | |
| *resident* | 73 | 78 | 25 | 31 | 0.47 (±0.19) | 0.70 | 0.24 | 0.48 |
| *5-10* | 53 | 43 | 21 | 13 | 0.57 (±0.19) | 0.80 | 0.33 | 0.55 |
| *15-20* | 29 | 24 | 8 | 6 | 0.55 (±0.24) | 0.83 | 0.25 | 0.55 |
| *20+* | 14 | 13 | 16 | 17 | 0.50 (±0.04) | 0.45 | 0.55 | 0.52 |
| Neuroradiologist | | | | | | | | |
| *5-10* | 16 | 16 | 19 | 16 | 0.52 (±0.02) | 0.50 | 0.54 | 0.50 |
| *10-15* | 11 | 12 | 21 | 19 | 0.51 (±0.11) | 0.37 | 0.64 | 0.48 |

| | | | | | | | | |
|---|---|---|---|---|---|---|---|---|
| Surgeon | | | | | | | | |
| *1-5* | 38 | 34 | 32 | 26 | 0.54 (±0.05) | 0.59 | 0.48 | 0.53 |
| *5-10* | 42 | 36 | 28 | 22 | 0.55 (±0.09) | 0.66 | 0.44 | 0.54 |
| General Practitioner | | | | | | | | |
| *1-5* | 28 | 29 | 5 | 4 | 0.50 (±0.30) | 0.87 | 0.15 | 0.49 |

Table H2. ORN detection performance results by specialty and years of experience of survey participants when using OPG images only.

| **Specialty (*years of experience*)** | **TP** | **FP** | **TN** | **FN** | **Accuracy (mean, SD)** | **Sensitivity** | **Specificity** | **Precision** |
|---|---|---|---|---|---|---|---|---|
| Dentist | | | | | | | | |
| *5-10* | 5 | 5 | 8 | 2 | 0.65 (±0.09) | 0.71 | 0.61 | 0.50 |
| *10-15* | 4 | 7 | 2 | 1 | 0.43 (±0.25) | 0.80 | 0.22 | 0.36 |
| *15-20* | 2 | 0 | 10 | 3 | 0.80 (±0.28) | 0.40 | 1.00 | 1.00 |
| Radiation Oncologist | | | | | | | | |
| *resident* | 3 | 11 | 3 | 4 | 0.29 (±0.10) | 0.43 | 0.21 | 0.21 |
| *5-10* | 6 | 13 | 7 | 4 | 0.43 (±0.13) | 0.60 | 0.35 | 0.31 |
| *15-20* | 3 | 6 | 2 | 2 | 0.39 (±0.15) | 0.60 | 0.25 | 0.33 |
| *20+* | 4 | 10 | 2 | 1 | 0.35 (±0.28) | 0.80 | 0.17 | 0.29 |
| Neuroradiologist | | | | | | | | |
| *5-10* | 1 | 5 | 6 | 3 | 0.47 (±0.18) | 0.25 | 0.54 | 0.17 |
| *10-15* | 3 | 8 | 3 | 2 | 0.37 (±0.15) | 0.60 | 0.27 | 0.27 |
| *20+* | 5 | 7 | 3 | 0 | 0.53 (±0.31) | 1.00 | 0.30 | 0.42 |
| Surgeon | | | | | | | | |
| *1-5* | 7 | 10 | 8 | 3 | 0.54 (±0.13) | 0.70 | 0.44 | 0.41 |
| *5-10* | 6 | 17 | 5 | 4 | 0.34 (±0.17) | 0.60 | 0.23 | 0.26 |
| General Practitioner | | | | | | | | |
| *1-5* | 4 | 6 | 3 | 1 | 0.50 (±0.21) | 0.80 | 0.33 | 0.40 |

Table H3. ORN detection performance results by specialty and years of experience of survey participants when using paired CT and OPG images.

| **Specialty (*years of experience*)** | **TP** | **FP** | **TN** | **FN** | **Accuracy (mean, SD)** | **Sensitivity** | **Specificity** | **Precision** |
|---|---|---|---|---|---|---|---|---|
| Dentist | | | | | | | | |
| *15-20* | 6 | 7 | 2 | 0 | 0.53 ±(0.09) | 1 | 0.22 | 0.46 |
| *10-15* | 4 | 4 | 4 | 0 | 0.67 ±(0.09) | 1 | 0.50 | 0.50 |
| General Practitioner | | | | | | | | |
| *1-5* | 5 | 7 | 1 | 0 | 0.46 ±(0) | 1 | 0.13 | 0.42 |
| Neuroradiologist | | | | | | | | |
| *5-10* | 4 | 1 | 9 | 0 | 0.93 ±(0.23) | 1 | 0.90 | 0.80 |
| *10-15* | 3 | 0 | 10 | 2 | 0.87 ±(0.23) | 0.60 | 1 | 1 |

| | | | | | | | | |
|---|---|---|---|---|---|---|---|---|
| *20+* | 4 | 7 | 3 | 0 | 0.50 ±(0.23) | 1 | 0.30 | 0.36 |
| Surgeon | | | | | | | | |
| *1 –5* | 10 | 2 | 16 | 0 | 0.93 ±(0.23) | 1 | 0.89 | 0.83 |
| *5-10* | 4 | 6 | 5 | 1 | 0.6 ±(0.23) | 1 | 0.45 | 0.4 |
| Radiation Oncologist | | | | | | | | |
| *resident* | 8 | 9 | 11 | 0 | 0.67 ±(0) | 1 | 0.55 | 0.47 |
| *5-10* | 5 | 4 | 4 | 0 | 0.69 ±(0.05) | 1 | 0.50 | 0.56 |
| *20+* | 4 | 2 | 7 | 1 | 0.79 ±(0.05) | 0.80 | 0.78 | 0.67 |

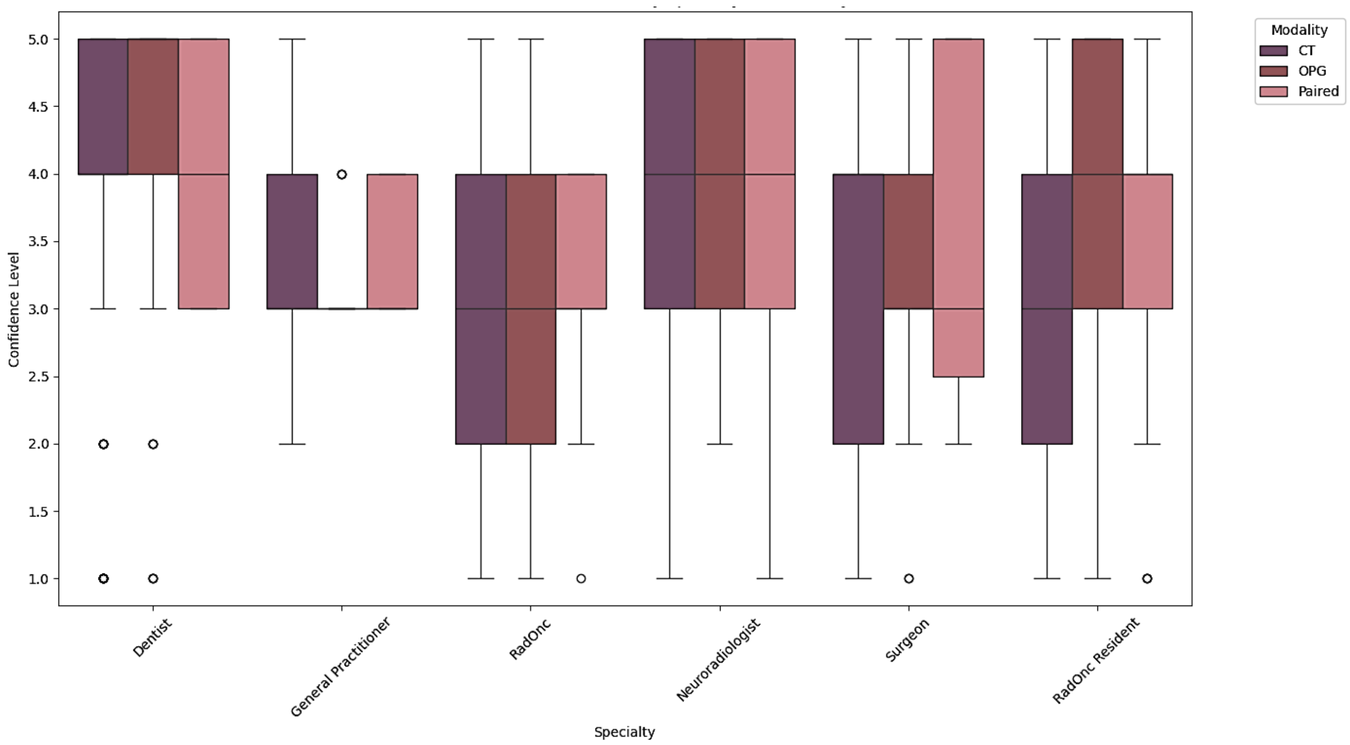


Figure H1. Distribution of level of confidence in detecting ORN by specialty and modality.

### Supplement I. DeLong Test results for ROC AUC Comparison

Table I1. Bonferroni-corrected p-values from DeLong's test comparing AUC values between different specialties for CT and OPG imaging alone.

| | Dentist | General Practitioner | Radiation Oncologist | Neuroradiologist | Surgeon | Radiation Oncologist Resident |
|---|---|---|---|---|---|---|
| **Dentist** | 0 | <0.001 | 1 | 1 | 1 | 0.01 |
| **General Practitioner** | <0.001 | 0 | <0.001 | <0.001 | <0.001 | 0.41 |
| **Radiation Oncologist** | 1 | <0.001 | 0 | 0.50 | 1 | 0.22 |
| **Neuroradiologist** | 1 | <0.001 | 0.50 | 0 | 1 | <0.001 |
| **Surgeon** | 1 | <0.001 | 1 | 1 | 0 | 0.019 |
| **Radiation Oncologist Resident** | 0.01 | 0.41 | 0.22 | <0.001 | 0.02 | 0 |

Table I2: AUC values for ORN detection by specialty, along with 95% confidence intervals (CI), p-values testing against the null hypothesis (AUC = 0.5), and Bonferroni-corrected p-values.

| Specialty | AUC | 95% CI Lower | 95% CI Upper | p-value | Bonferroni Corrected p-value |
|---|---|---|---|---|---|
| Dentist | 0.55 | 0.49 | 0.61 | 0.13 | 0.80 |
| General Practitioner | 0.5 | 0.39 | 0.61 | 1 | 1 |
| Radiation Oncologist | 0.55 | 0.50 | 0.61 | 0.06 | 0.37 |
| Neuroradiologist | 0.5 | 0.43 | 0.58 | 1 | 1 |
| Surgeon | 0.52 | 0.46 | 0.57 | 0.50 | 1 |
| Radiation Oncologist Resident | 0.46 | 0.39 | 0.52 | 0.18 | 1 |

Table I3. Bonferroni-corrected p-values from DeLong's test comparing AUC values between different specialties for paired 2D/3D imaging.

| | Dentist | Residents | Neuroradiologist | Surgeon | RadOnc |
|---|---|---|---|---|---|
| **Dentist** | 0 | 1 | 0.92 | 1 | 1 |
| **Residents** | 1 | 0 | 0.56 | 1 | 1 |
| **Neuroradiologist** | 0.92 | 0.56 | 0 | 1 | 1 |
| **Surgeon** | 1 | 1 | 1 | 0 | 1 |
| **Radiation Oncologist** | 1 | 1 | 1 | 1 | 0 |

Table I4: AUC values for ORN detection by specialty, along with 95% confidence intervals (CI), p-values testing against the null hypothesis (AUC = 0.5), and Bonferroni-corrected p-values for paired 2D/3D imaging evaluation.

| Specialty | AUC | 95% CI Lower | 95% CI Upper | p-value | Bonferroni Corrected p-value |
|---|---|---|---|---|---|
| Dentist | 0.88 | 0.72 | 1.03 | <0.001 | <0.001 |
| Residents | 0.79 | 0.64 | 0.93 | <0.001 | <0.001 |
| Neuroradiologist | 0.85 | 0.73 | 0.97 | <0.001 | <0.001 |
| Surgeon | 0.98 | 0.93 | 1.03 | 0 | 0 |
| Radiation Oncologist | 0.82 | 0.69 | 0.95 | <0.001 | <0.001 |

### Supplement J. Confidence in ORN staging analyses

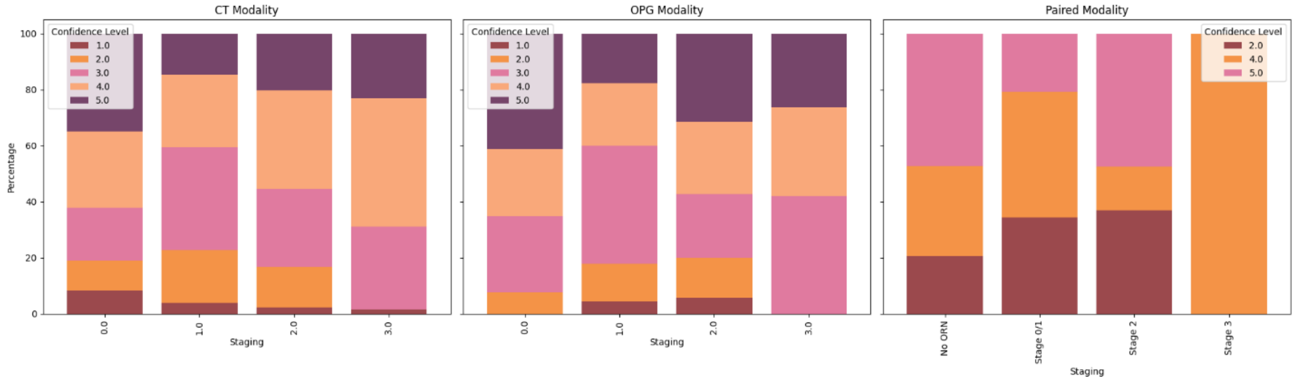


Figure J1. Distribution of level of confidence in staging ORN across all specialties when using CT, OPG and paired images.
